# Seroprevalence of syphilis among sex workers in Nairobi, Kenya: a retrospective study

**DOI:** 10.64898/2026.03.22.26349033

**Authors:** Kristi Loeb, Alice van Wyk, Kieran Milner, Candice Lemaille, Christina Frederick, Mikayla Hunter, Brielle Martens, Julie Lajoie, Placide Mbala-Kingebeni, Anne W. Rimoin, Nicole A. Hoff, Ryan S. Noyce, Keith R Fowke, Joshua Kimani, Lyle R. McKinnon, Souradet Y. Shaw, Derek R. Stein, Jason Kindrachuk

**Affiliations:** Department of Medical Microbiology & Infectious Diseases, Max Rady College of Medicine, University of Manitoba, Winnipeg, Manitoba, Canada; Cadham Provincial Laboratory, Winnipeg, MB, Canada; College of Community and Global Health, University of Manitoba, Winnipeg, Manitoba, Canada; Department of Epidemiology, UCLA Fielding School of Public Health, Los Angeles, California, United States; Pathogen Genomics Laboratory, Epidemiology and Global Health Department, Institut National de Recherche Biomédicale, Kinshasa, Democratic Republic of the Congo; Service de Microbiologie, Cliniques Universitaires, Faculté de Médecine, Université de Kinshasa, Kinshasa, Democratic Republic of the Congo; Department of Medical Microbiology and Immunology, University of Alberta, Edmonton, Canada; The Li Ka Shing Institute of Virology, University of Alberta, Edmonton, Canada; Department of Medical Microbiology, University of Nairobi, Nairobi, Kenya; National Microbiology Laboratory Branch, Public Health Agency of Canada, Winnipeg, Canada

## Abstract

Syphilis is a sexually transmitted and bloodborne infection caused by *Treponema pallidum* spp. *pallidum*. Given the paucity of data on syphilis in Kenyan sex workers and gay, bisexual, and other men who have sex with men (GBMSM), we conducted a retrospective study of syphilis seropositivity in female sex workers (FSW) and GBMSM in Nairobi, Kenya. Seropositivity testing of cryopreserved plasma samples showed that 11.1% (72/647) were positive. Syphilis seropositivity was associated with HIV status, and FSWs were disproportionately represented in the seropositive group (66/72, 92%). Here, we report a higher seropositive rate than in previous studies in Kenya, and ongoing community and surveillance supports are important for addressing the ongoing public health impacts of syphilis.

## Introduction

Syphilis is a sexually transmitted and bloodborne infection (STBBI) caused by *Treponema pallidum* spp. *pallidum*. When untreated, syphilis can eventually lead to several complications, including congenital syphilis in pregnant people, cardiovascular lesions, and dementia [1]. Though prevention, testing, and treatment are well-established, syphilis continues to be a significant cause of global morbidity and mortality. In 2022, there were an estimated 8 million new cases of infectious syphilis in adults and 700,000 cases of congenital syphilis globally [2]. Low- and middle-income countries (LMICs) are disproportionately affected, with rates of both infectious and congenital syphilis recently increasing across Africa, and the highest estimated rates of congenital syphilis cases in Western and Central Africa, followed by Eastern and Southern Africa [2]. Despite this, comprehensive surveillance for syphilis is lacking in the most-affected countries. Surveillance that is conducted is generally done on antenatal populations, given the burden of congenital syphilis; however, this brings the risks of missing syphilis in other high-risk populations.

Syphilis frequently overlaps with HIV and other STBBIs, with syphilis, HIV, and other STBBIs disproportionately affecting high-risk communities. In particular, female sex workers (FSWs), male sex workers, and gay, bisexual, and other men who have sex with men (GBMSM) are especially affected by STBBIs [3, 4]. These communities are also profoundly affected by stigma, whether related to their work, sexual orientation, gender, or illness [5]. Given the paucity of data on syphilis in Kenyan sex workers and GBMSM, we conducted a retrospective study of syphilis seropositivity in FSWs and GBMSM in Nairobi, Kenya.

## The Study

We analyzed cryopreserved plasma samples from participants of the Sex Worker Outreach Program (SWOP) in Nairobi, Kenya [6]. Samples were randomly selected from the available samples within the SWOP cohort, which were originally collected between 2013 and 2019 during previous studies with two participating labs and then cryopreserved (Table 1). This study’s cohort was primarily composed of individuals who identified as FSWs (563/647) and a smaller proportion of male sex workers (MSWs), all of whom also identified as GBMSM (77/647). The majority of the study’s cohort included people living with HIV (PLWH) (555/647). Participants ranged in age from 19 to 56 at the time of sampling (Table 1). Undiluted samples were tested for syphilis using the Abbott ARCHITECT syphilis TP chemiluminescent microparticle immunoassay according to the manufacturer’s instructions. Samples with treponemal antibody reactivity ≥1.00 were considered positive. Descriptive statistical and chi-square analysis were performed to assess for associations.

**Table 1.** Demographic data for study participants. Percentages of row totals are presented in parentheses. Samples with unknown qualities were omitted from statistical analysis.

|  | Positive | Negative | Total |
| --- | --- | --- | --- |
| **Age group** |  |  |  |
| ≤30 | 3 (7.0) | 40 (93) | **43** |
| 31-44 | 14 (5.3) | 252 (95) | **266** |
| 45-54 | 36 (32) | 76 (67) | **112** |
| ≥55 | 12 (92) | 1 (7.7) | **13** |
| Unknown | 7 (3.3) | 206 (97) | **213** |
| **Total** | **72** | **575** | **647** |
|  |  |  | **p<0.0001** |
| **Sample collection year** |  |  |  |
| 2013-16 | 4 (6.2) | 61 (94) | **65** |
| 2017-19 | 62 (12) | 467 (88) | **529** |
| Unknown | 6 (11) | 47 (89) | **53** |
| **Total** | **72** | **575** | **647** |
|  |  |  | **p=0.1348** |
| **HIV status** |  |  |  |
| Negative | 1 (1.2) | 84 (99) | **85** |
| Positive | 70 (13) | 485 (87) | **555** |
| Unknown | 1 (14) | 6 (86) | **7** |
| **Total** | **72** | **575** | **647** |
|  |  |  | **p=0.0018** |
| **Key population** |  |  |  |
| Female sex worker | 66 (12) | 497 (88) | **563** |
| GBMSM/Male sex worker | 1 (1.3) | 76 (99) | **77** |
| Unknown | 5 (71) | 2 (29) | **7** |
| **Total** | **72** | **575** | **647** |
|  |  |  | **p=0.0051** |
|  |  | **Negative** | **Total** |

Out of the 647 samples tested, a total of 72 (11.1%) were seropositive for syphilis. Making up half of the positive samples (36/72); the 45-54 age group was significantly overrepresented in the seropositive group (p<0.0001), though this age group also made up the largest proportion of the cohort (266/647, 41.1%). Seropositivity was next highest in the 31-44 year olds (14/72, 19.4%) and closely followed by ≥55 year-olds (12/72, 16.6%). HIV status was associated with syphilis seropositivity; syphilis seropositivity was significantly higher in PLWH (p=0.0018). FSWs were disproportionately represented in the seropositive group (66/72, 92%), with an 11.2% seropositivity rate in the FSW group compared to only 1.3% in MSWs (p=0.0051), though it is worth noting there was a relatively small sample size for both MSWs compared to FSWs (77 to 563).

## Conclusion

Here, we report a higher seropositive rate than in previous studies in Kenya [7, 8]. As the test does not differentiate between active or prior infection, the seropositivity in the 45-54 and ≥55 age groups may reflect the increased amount of time in sex work, leading to more potential syphilis exposures as compared to a younger person. There may also be different habits related to condom use that may affect the risk of sexual behaviours between age groups [9]. The age-adjusted disability-adjusted life years (DALYs) per 100,000 population due to syphilis in Kenya has recently been estimated as 177.7, indicating a burden about tenfold below the leading overall causes of morbidity and mortality in Kenya [8, 10]. Our data suggest there may be a particularly high burden among high-risk groups. The high rate of syphilis positivity in FSWs is particularly concerning, given the possibility of congenital syphilis. For women who become pregnant, this can lead to serious complications, including low birth weight, premature delivery, and stillbirth. Though screening may be available, it is not always accessible and does not necessarily prevent adverse neonatal outcomes [7]. As of 2023, 93.6% of women who accessed antenatal care in Kenya were tested for syphilis, and only 0.95% were reported to have tested positive, and only 44.3% of those women received treatment, which is well below the WHO’s goal of ≥95%, leaving a sizeable gap that increases the potential risk of congenital syphilis [2]. As of 2022, Kenya only recommends single antenatal syphilis screening, further risking undetected maternal syphilis and the opportunity for congenital syphilis to occur [7]. Beyond the risk of congenital syphilis, untreated syphilis can also result in long-term consequences for the individuals themselves, such as neurosyphilis, with risks increased for PLWH [11].

We previously showed probable mpox exposure within the same samples from this cohort [12]. Alongside other studies that have demonstrated mpox transmission within sexual networks, including within Kenya [13], the association of syphilis and HIV positivity found here further demonstrates an urgent need for comprehensive STBBI surveillance in Kenya and adjacent regions. The ongoing Clade Ib mpox outbreak has highlighted the sex worker community as being critically vulnerable to both the outbreak itself and a public health response that is not tailored to their unique needs [14]. Our data further indicate a potential overlap between two STBBIs that can each have devastating consequences when left untreated. We report a significantly higher seroprevalence of syphilis than previously identified in the general population, in a population largely composed of PLWH. It is important to note that this study was conducted using a convenience sample and may not be generalizable to larger populations. Further studies must be conducted in these key populations to gain better insight into the true burden of disease. We were additionally limited by the inability to differentiate between historical and current syphilis infection. Future studies are needed to identify active infection and define the prevalence within these populations. Conducting surveillance and providing support to these communities are important steps in addressing the ongoing burden due to syphilis.

**Figure 1.**
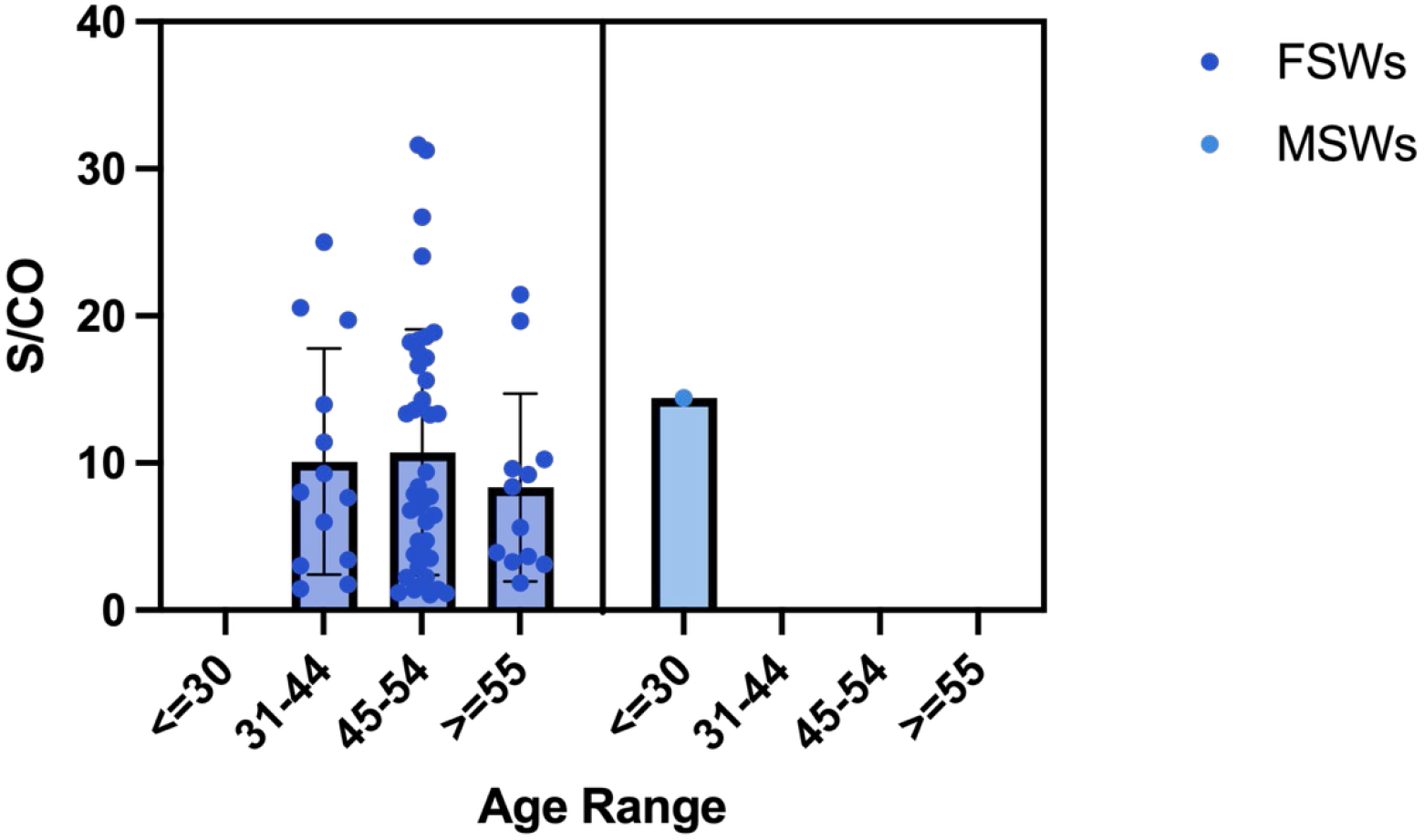
Retrospective analysis of syphilis seropositivity within sex workers in Kenya. Seropositivity was detected in 71 FSWs, 1 GBMSM/MSW, and 3 community members. An S/CO of 1.00 was used for positivity (negative samples not shown). Horizontal bars indicate standard deviation.

## Data Availability

All data produced in the present study are available upon reasonable request to the authors.

## Acknowledgements

We are grateful to the Kenyan study participants for their longstanding involvement in our research projects. This work was supported through funding from the Canadian Institutes of Health Research and International Development Research Centre (Grant No. 202209MRR-489062-MPX-CDAA-168421), a Canadian Institutes of Health Research Tier 2 Canada Research Chair (Grant Number 950-231498 to J.K.), and Department of Defense, Defense Threat Reduction Agency, Monkeypox Threat Reduction Network (MPX-TRN) grant #HDTRA1-21-1-0040 and United States Department of Agriculture Non-Assistance Cooperative Agreement #20230048.

## Notes

### Competing Interest Statement

The authors have declared no competing interest.

### Author Declarations

University of Manitoba Research Ethics Board approval was obtained during the original sample collection. For this study, previously collected historical samples (cryopreserved plasma) were utilized for all analyses.

## References

1. Peeling, R.W., et al., Syphilis. The Lancet, 2023. 402(10398): p. 336–346.

2. World Health Organization, Data on syphilis. 2025: The Global Health Observatory: Sexually Transmitted Infections.

3. Gedfie, S., et al., Prevalence and associated factors of syphilis among female sex workers in East Africa: a systematic review and meta-analysis. Front Public Health, 2025. 13: p. 1543119.

4. Smith, A.D., et al., HIV burden and correlates of infection among transfeminine people and cisgender men who have sex with men in Nairobi, Kenya: an observational study. Lancet HIV, 2021. 8(5): p. e274–e283.

5. Babu, H., et al., Trends in HIV Incidence among Female Sex Workers and Men who Have Sex with Men in Nairobi, Kenya, 2009-2021. AIDS Behav, 2025. 29(7): p. 2226–2233.

6. Omollo, K., et al., Differential Elevation of Inflammation and CD4(+) T Cell Activation in Kenyan Female Sex Workers and Non-Sex Workers Using Depot-Medroxyprogesterone Acetate. Front Immunol, 2020. 11: p. 598307.

7. Laktabai, J., et al., Associations between Antenatal Syphilis Test Results and Adverse Pregnancy Outcomes in Western Kenya. Am J Trop Med Hyg, 2022. 107(2): p. 401–406.

8. Yu, W., X. You, and W. Luo, Global, regional, and national burden of syphilis, 1990-2021 and predictions by Bayesian age-period-cohort analysis: a systematic analysis for the global burden of disease study 2021. Front Med (Lausanne), 2024. 11: p. 1448841.

9. Parcesepe, A.M., et al., Early sex work initiation and condom use among alcohol-using female sex workers in Mombasa, Kenya: a cross-sectional analysis. Sex Transm Infect, 2016. 92(8): p. 593–598.

10. World Health Organization, Disease Burden 2000-2021, in The Global Health Observatory. 2026.

11. Njiru, E., et al., Early neurosyphilis presenting with facial palsy and an oral ulcer in a patient who is human immunodeficiency virus positive: a case report. J Med Case Rep, 2017. 11(1): p. 134.

12. Loeb, K., et al., Retrospective Seroprevalence of Orthopoxvirus Antibodies among Key Populations, Kenya. Emerg Infect Dis, 2024. 30(9): p. 1944–1947.

13. Mutuku, P., et al., Clade Ib Mpox Outbreak - Kenya, July 2024-February 2025. MMWR Morb Mortal Wkly Rep, 2025. 74(22): p. 379–384.

14. Adebisi, Y.A., et al., Sex Workers and the Mpox Response in Africa. J Infect Dis, 2024. 230(4): p. 786–788.

